# Analysis of the Second COVID-19 Wave in India Using a Birth-Death Model

**DOI:** 10.1101/2021.05.19.21257447

**Authors:** Narayanan C. Viswanath

## Abstract

India is witnessing the second wave of the COVID-19 disease from the first half of February 2021. The method in [5] is applied here to analyze the second wave in India. We start with fitting a birth-death model to the active and total cases data for the period from 13^th^ to 28^th^ February 2021. This initial dataset is expanded step by step by adding the two future week’s data to it until 14^th^ May 2021. This resulted in six models in total. The efficacy of each model is tested in terms of predictions made for the next two weeks. The infectivity rates are found to be ever-increasing in the case of the five initial models. The infectivity rate for the sixth model, which is based on the data from 13^th^ February to 14^th^ May 2021, shows a decreasing nature with an increase in time. This indicates a decline in the second wave, which may start from 4^th^ June 2021 according to the fitted parameters.

## Introduction

According to Ranjan et al. [3], a rise in the number of COVID-19 cases in India started from 13^th^ February 2021 which indicated the start of the second wave of the disease progression. Using a SIR model [2], they predict a peaking of the same in mid-May 2021.

[4] applied a generalized birth-death model [1] for modeling the total and actively infected COVID-19 cases in several countries. This turned out to be a special case, of the standard SIR model [2], in which the susceptible cases variable has no explicit role. [5] discussed application of the model in [4] when a rise in the number of cases occurs after a fall. Here we check the efficacy of the method in [5] in analyzing the second wave that occurred in India.

## Methods

By taking the infection birth rate as *λ*(*t*) = *ae*^−*bt*^ and the recovery rate as *µ* in a generalized birth-death model [1], [4] derives the following equations for the actively infected *I*(*t*) and total *M*(*t*) cases as:

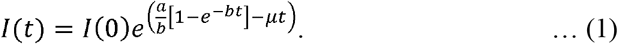

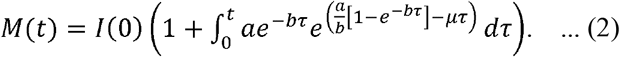

[5] discussed application of the above model when a rise in the number of cases occurs after a fall. Their idea was to fit equations (1) and (2) to a dataset after adjusting the total cases so that the total and active cases remain the same at the starting epoch. We follow the method in [5] for fitting equations (1) and (2) to the active and total cases data [7]. Fitting was done using the *nlinfit* function available in the MATLAB R2019b [6] software.

## Results and Discussion

We consider 13^th^ February 2021 as a starting point. We fitted equations (1) and (2) using the method discussed in [5] for the data from 13^th^ to 28^th^ February 2021. Figure 1 shows the fit. The parameters for this fit are: a = 0.0794821535; b = -0.0157707070; *I*(0) = 132167; *µ* = 0.0739635918. Here notice that the negative value for parameter *b* indicates that the infection rate is going to increase forever. This according to us points to a failure of the model as far as a long-term prediction is concerned. The absence of the number of susceptible cases in the model could be the reason for this phenomenon. Figure 2 is the comparison of the predictions by the fitted model with actual data for the next two weeks (from 1^st^ to 14^th^ March 2021). Figure 2 shows that the accuracy of the prediction decreases as time increases.

**Figure 1.**
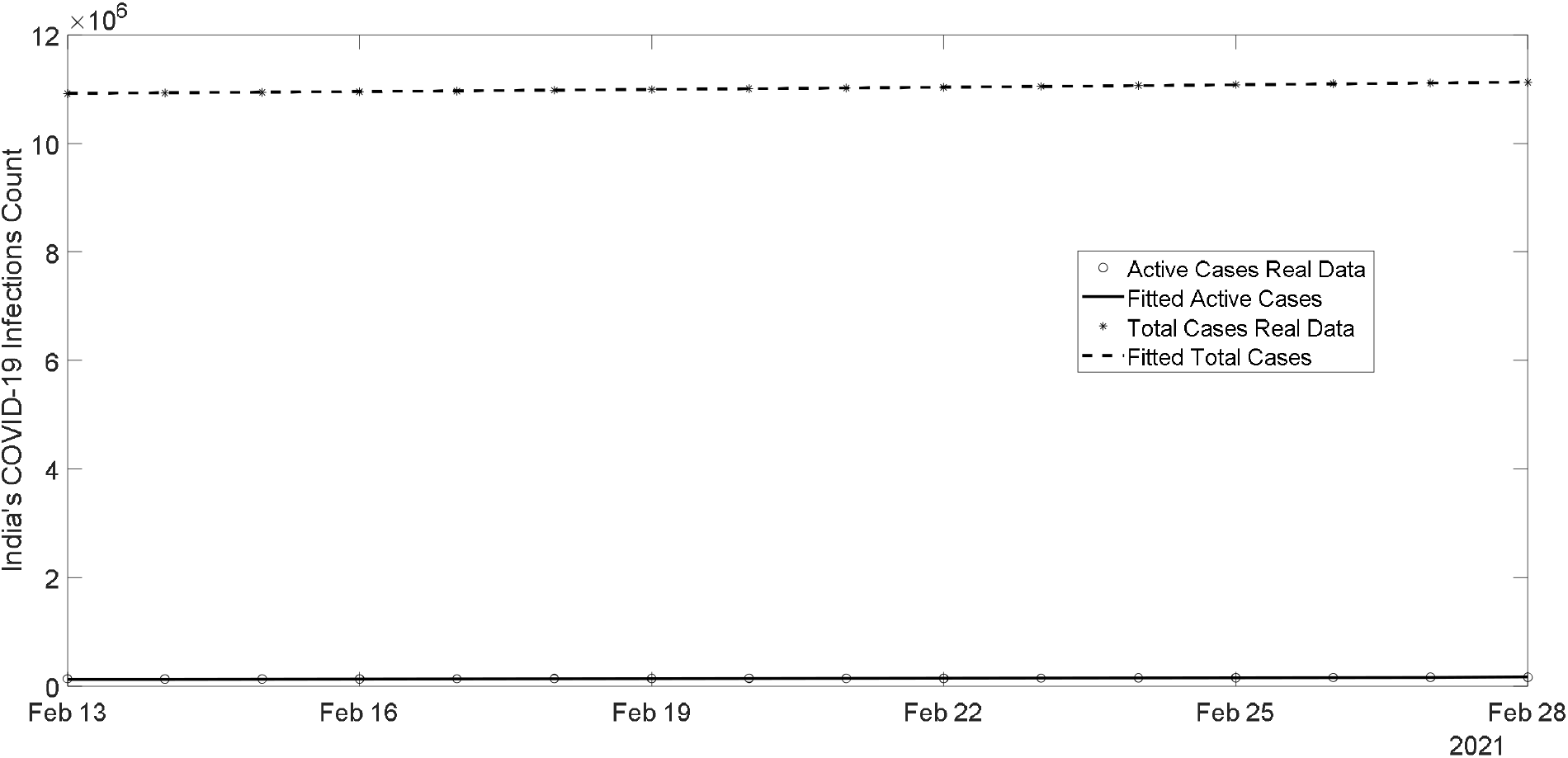
Comparison of the fitted values of active and total cases with the actual values for India based on the data from 13^th^ to 28^th^ February 2021. The parameters for this fit are: a = 0.0794821535; b = -0.0157707070; *I*(0) = 132167; *µ* = 0.0739635918.

**Figure 2.**
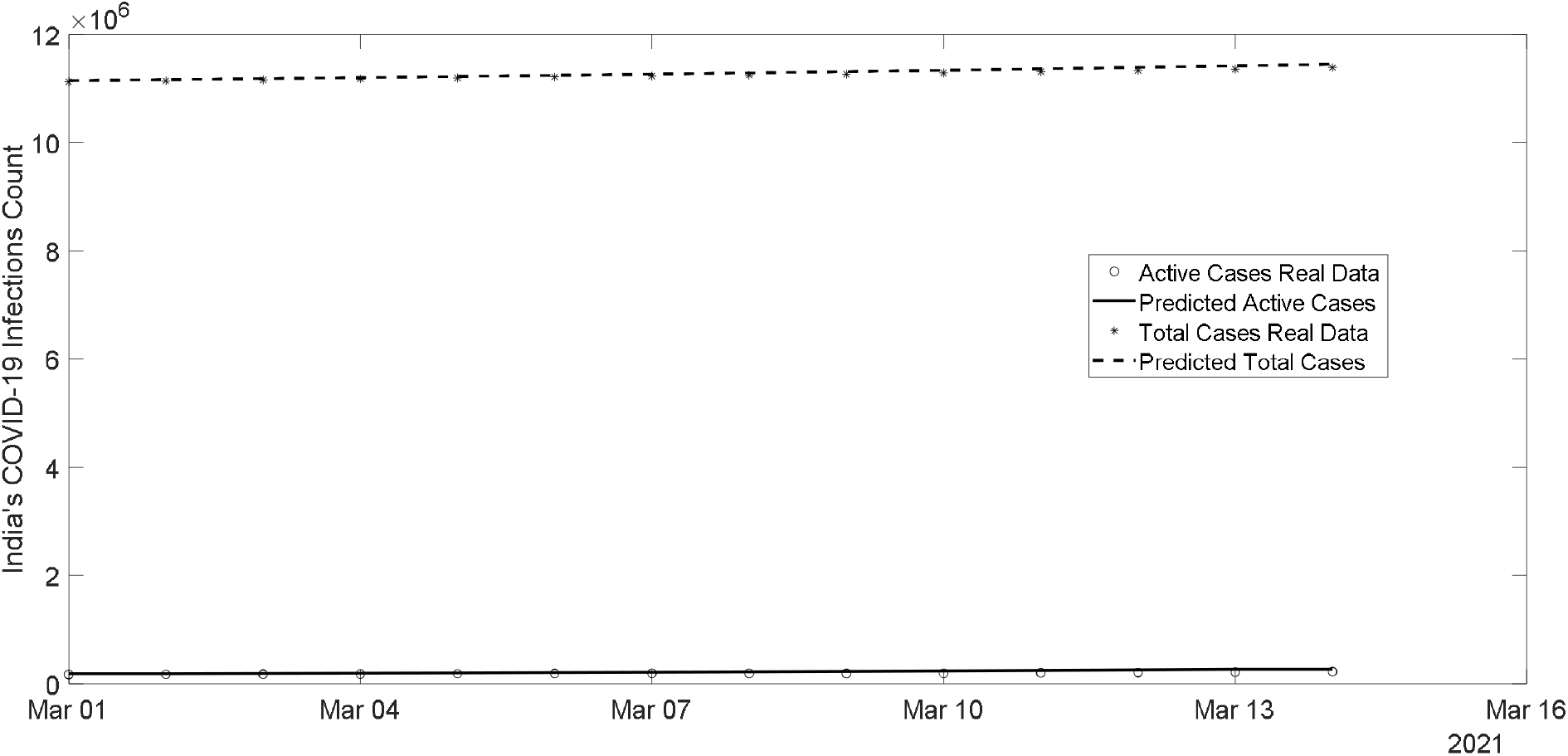
Comparison of the predicted active and total cases with their actual values for India from 1^st^ to 14^th^ March 2021, when fitting was done based on the data from 13^th^ to 28^th^ February 2021.

Next, we expanded the prediction dataset by adding the actual data of the next two weeks to it. Figure 3 shows the fit for the data from 13^th^ February to 14^th^ March 2021. The parameters for this fit are: a = 0.0895802716; b = -0.0046661410; *I*(0) = 128946; *µ* = 0.0782528444. Again the parameter *b* turns out to be negative. Prediction for the next two weeks, which is from 15^th^ to 28^th^ March 2021, is given in Figure 4. It shows a decent prediction, whose accuracy begins to decrease as time increases.

**Figure 3.**
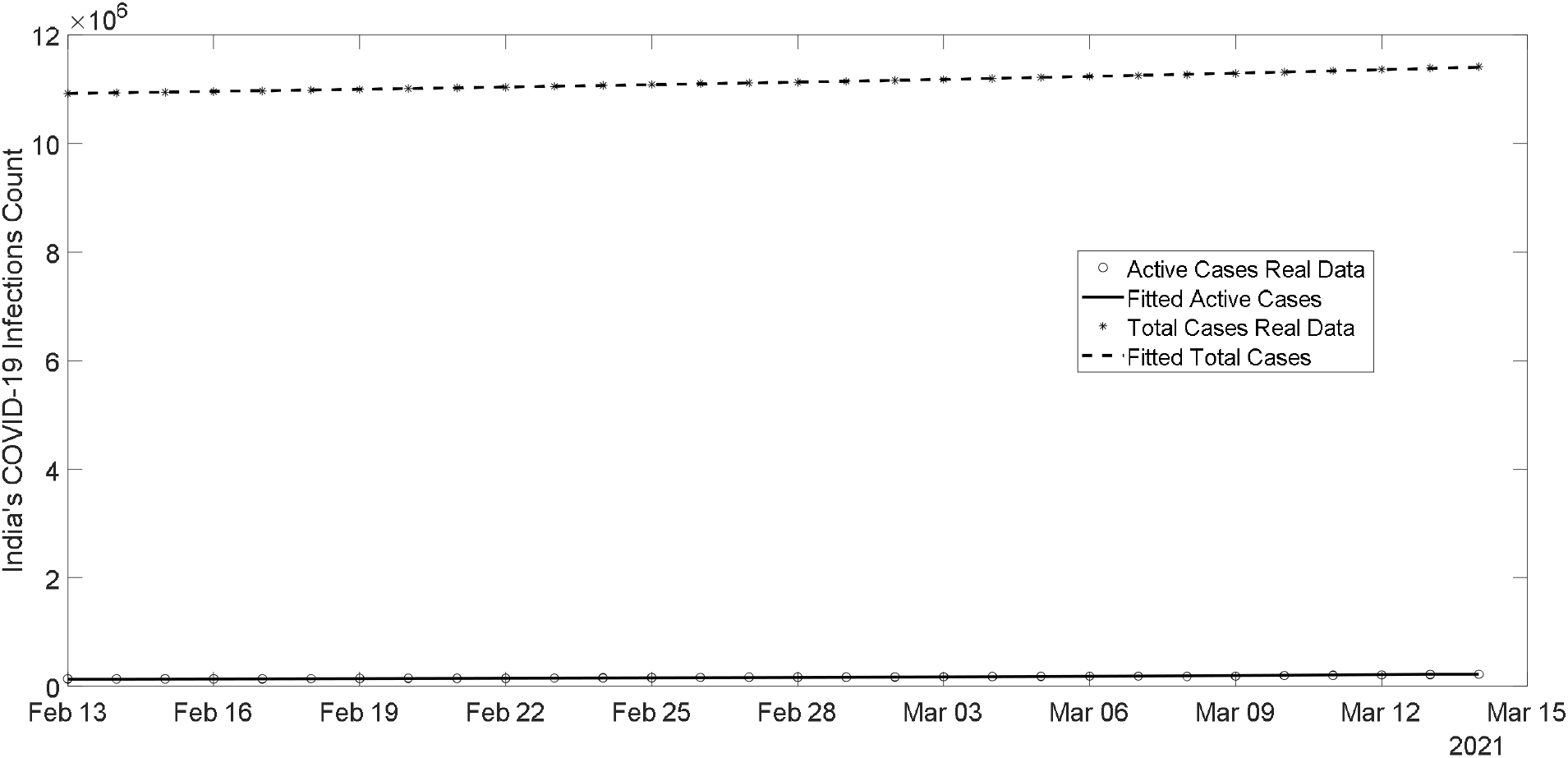
Comparison of the fitted values of active and total cases with the actual values for India based on the data from 13^th^ February to 14^th^ March 2021. The parameters for this fit are: a = 0.0895802716; b = -0.0046661410; *I*(0) = 128946; *µ* = 0.0782528444.

**Figure 4.**
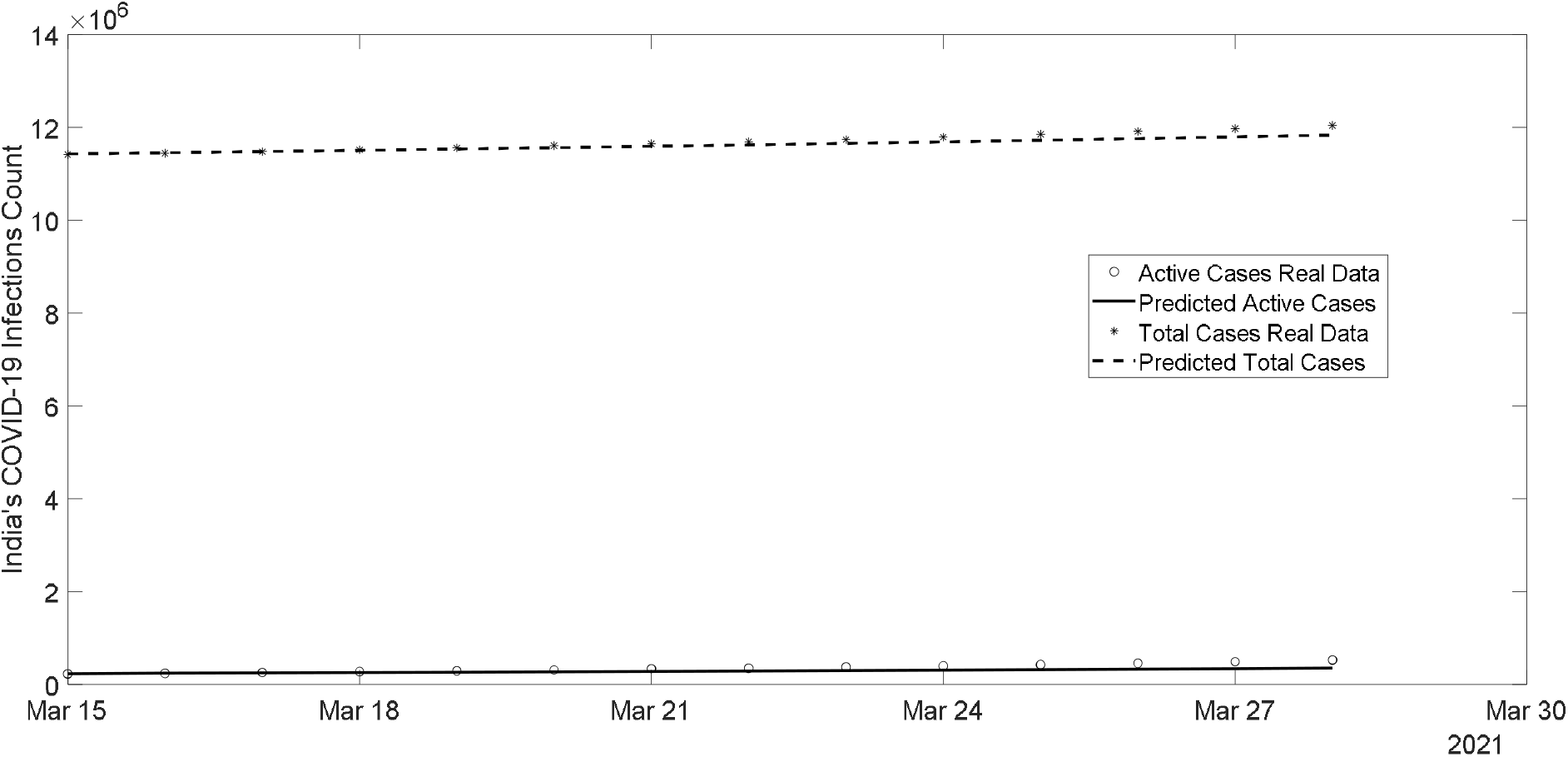
Comparison of the predicted active and total cases with their actual values for India from 15^th^ to 28^th^ March 2021, when fitting was done based on the data from 13^th^ February to 14^th^ March 2021.

We then considered the data from 13^th^ February to 31^st^ March 2021. Figure 5 shows the fit whose parameters are given by: a = 0.0784394479; b = -0.0135156063; *I*(0) = 135863; *µ* = 0.0778148298. Figure 6 shows the predictions for the next two weeks: from 1^st^ to 14^th^ April 2021. The parameters for Figure 7, which shows the fit for the data from 13^th^ February to 14^th^ April 2021, are given by: a = 0.0868287781; b = - 0.0090513907; *I*(0) = 122037; *µ* = 0.0750109712. Figure 8 shows the quality of predictions for the next two weeks (from 15^th^ to 28^th^ April 2021). Figures 5 to 8 reveal that while the quality of fit (Figures 5 and 7) remains the same, the quality of future prediction decreases (Figures 6 and 8).

**Figure 5.**
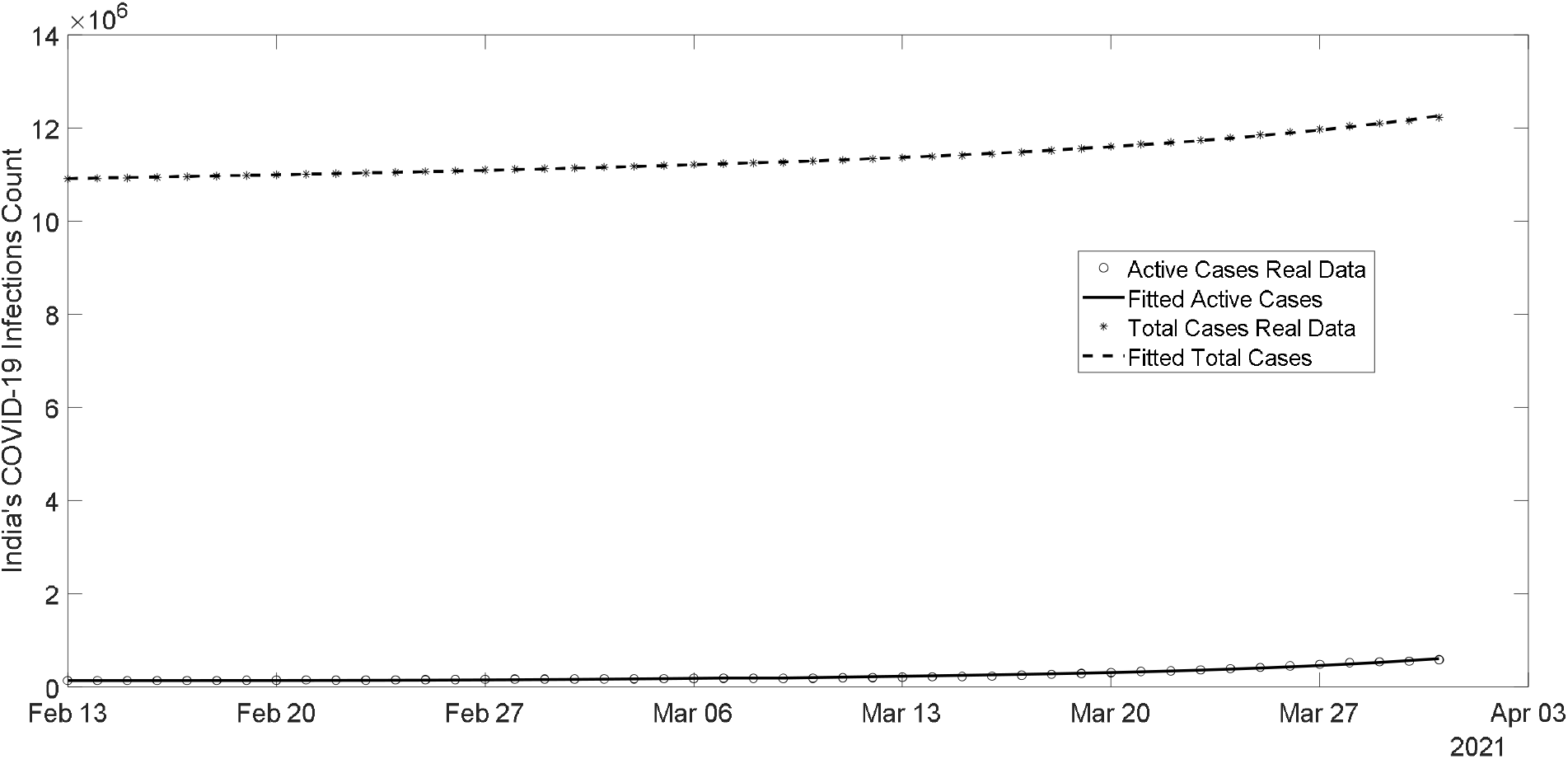
Comparison of the fitted values of active and total cases with the actual values for India based on the data from 13^th^ February to 31^st^ March 2021. The parameters for this fit are: a = 0.0784394479; b = -0.0135156063; *I*(0) = 135863; *µ* = 0.0778148298.

**Figure 6.**
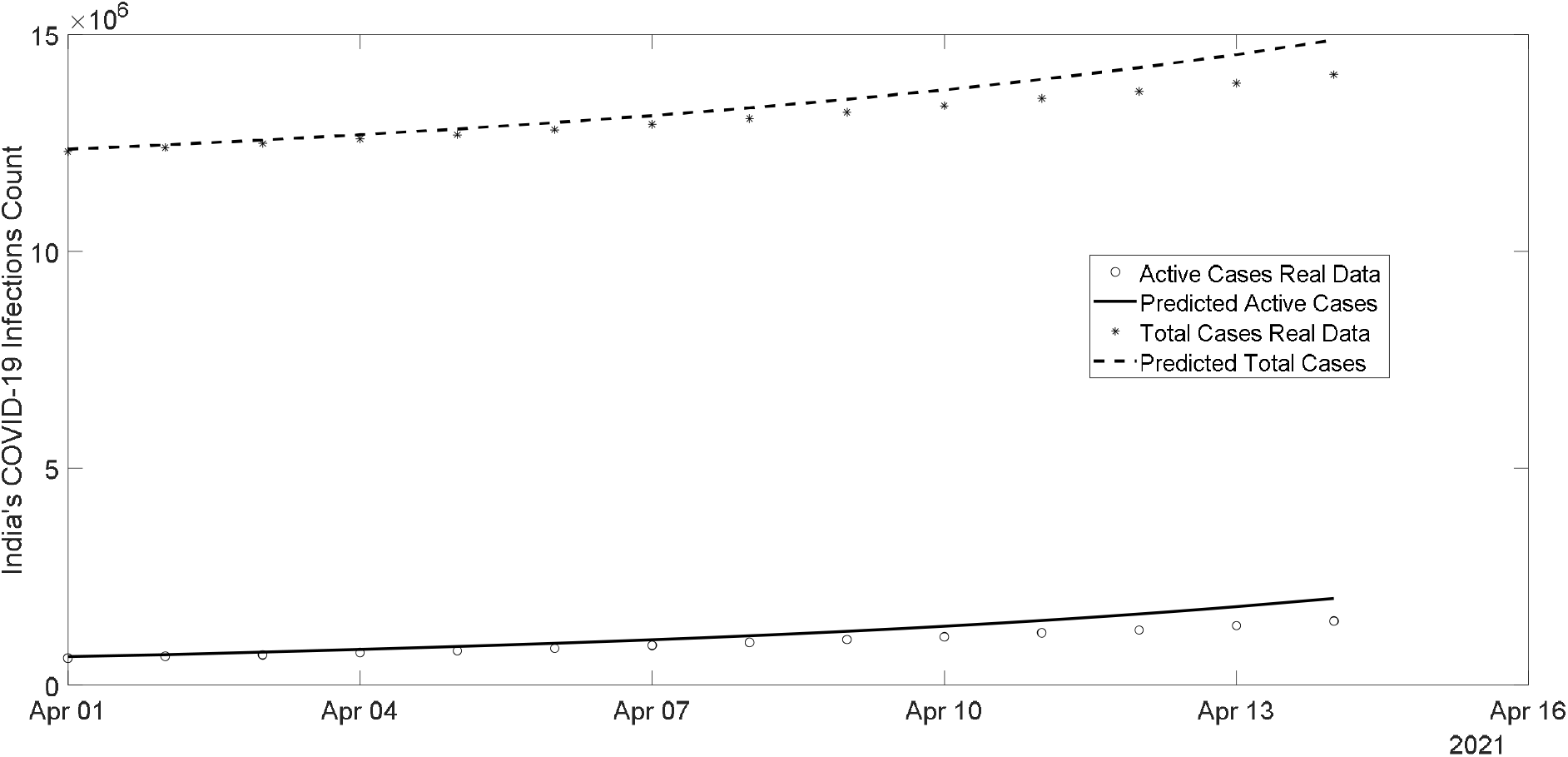
Comparison of the predicted active and total cases with their actual values for India from 1^st^ to 14^th^ April 2021, when fitting was done based on the data from 13^th^ February to 31^st^ March 2021.

**Figure 7.**
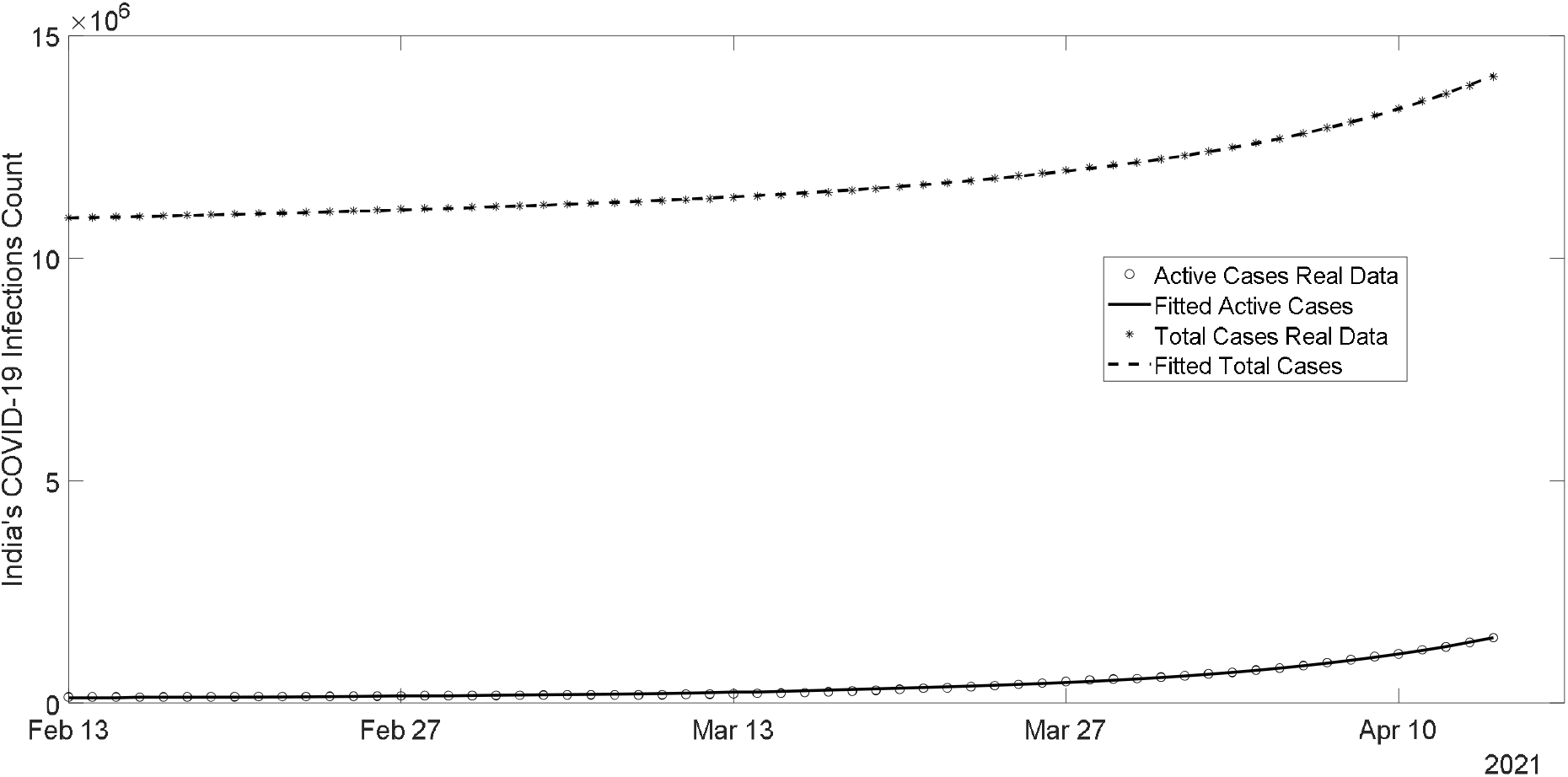
Comparison of the fitted values of active and total cases with the actual values for India based on the data from 13^th^ February to 14^th^ April 2021. The parameters for this fit are: a = 0.0868287781; b = - 0.0090513907; *I*(0) = 122037; *µ* = 0.0750109712.

**Figure 8.**
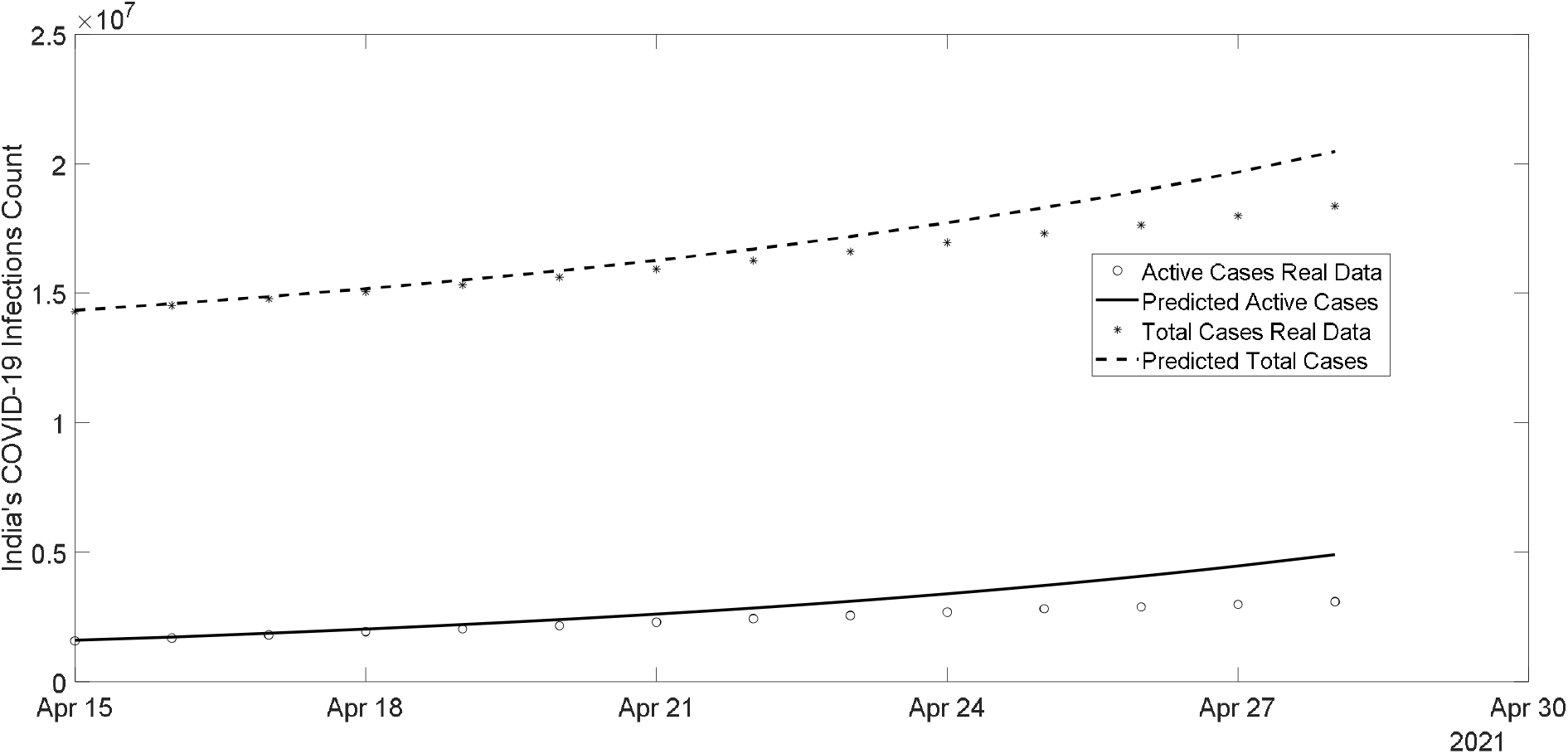
Comparison of the predicted active and total cases with their actual values for India from 15^th^ to 28^th^ April 2021, when fitting was done based on the data from 13^th^ February to 14^th^ April 2021.

Figure 9 shows that the fitted counts are slightly greater than the actual values towards the end of the fit, when fitting was done on the data from 13^th^ February to 30^th^ April 2021. The parameters for this fit are: a = 0.12608518091; b = - 0.00129869234; *I*(0) = 58896; *µ* = 0.07907911057. The predicted values for the period of 1^st^ to 14^th^ of May 2021 are much higher than the actual values as can be observed from Figure 10. More specifically, the actual and predicted active cases for May 14^th^ are 3679691 and 8594118 respectively. The actual and predicted total cases for the same date are 24372243 and 30651576 respectively.

**Figure 9.**
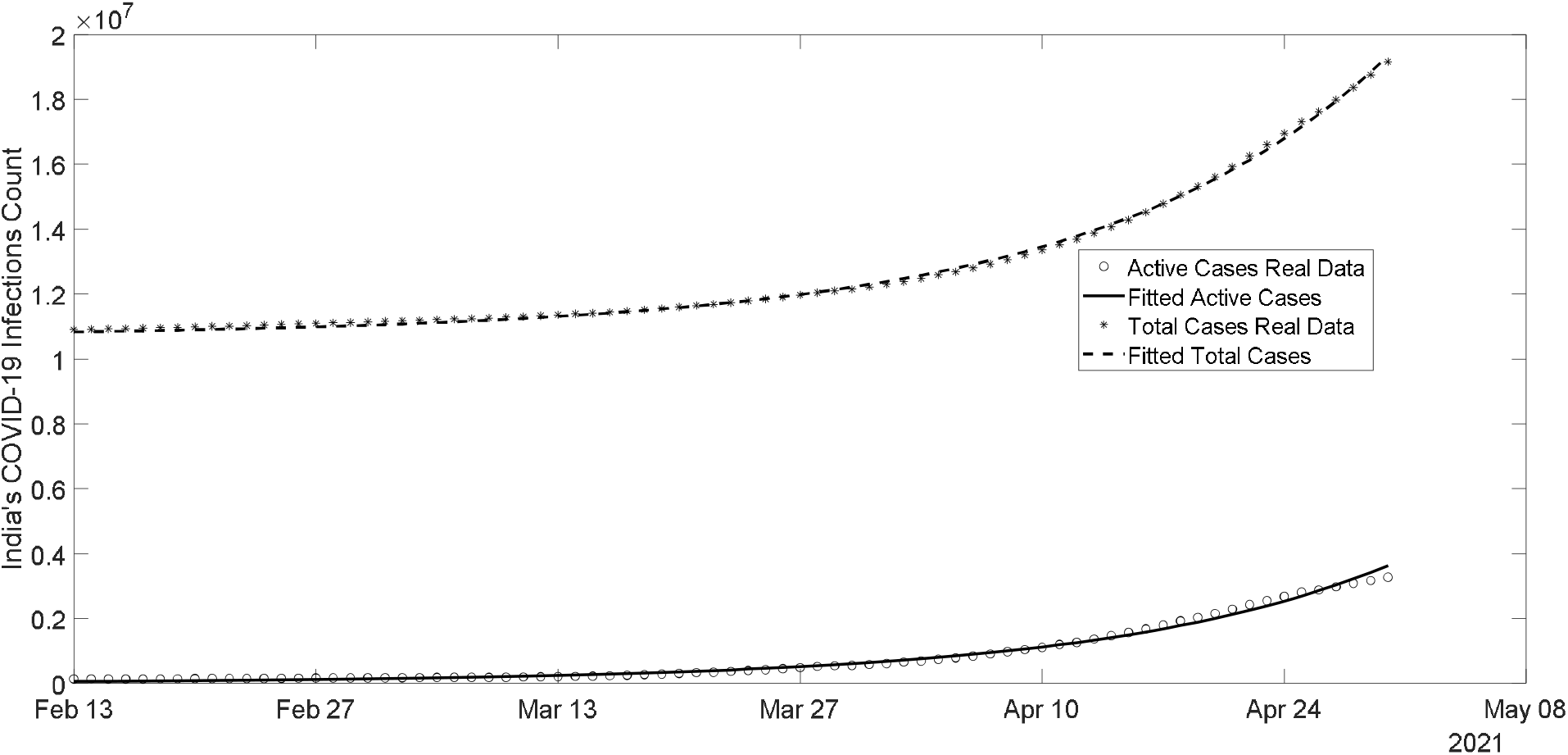
Comparison of the fitted values of active and total cases with the actual values for India based on the data from 13^th^ February to 30^th^ April 2021. The parameters for this fit are: a = 0.12608518091; b = - 0.00129869234; *I*(0) = 58896; *µ* = 0.07907911057.

**Figure 10.**
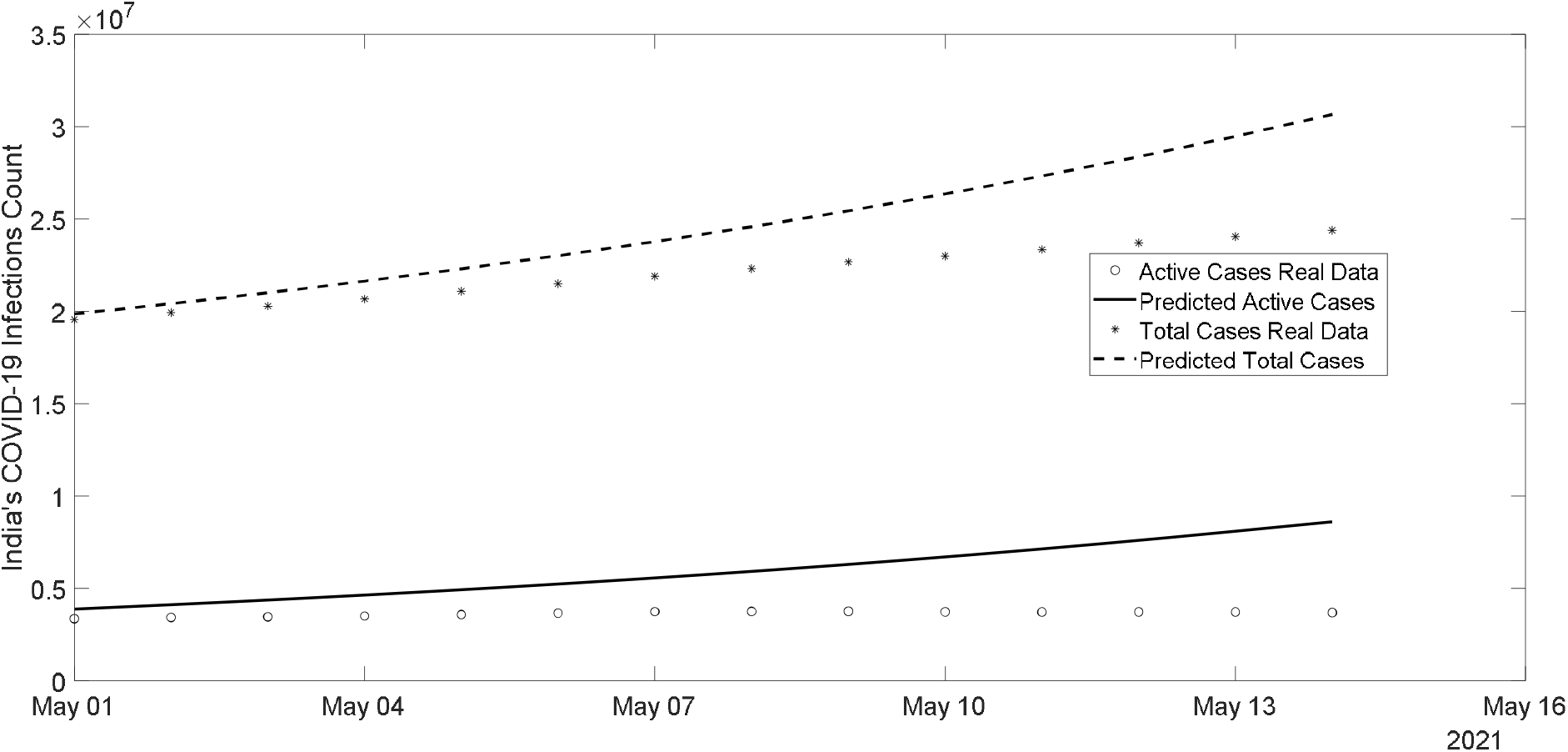
Comparison of the predicted active and total cases with their actual values for India from 1^st^ to 14^th^ May 2021, when fitting was done based on the data from 13^th^ February to 30^th^ April 2021.

**Figure 11.**
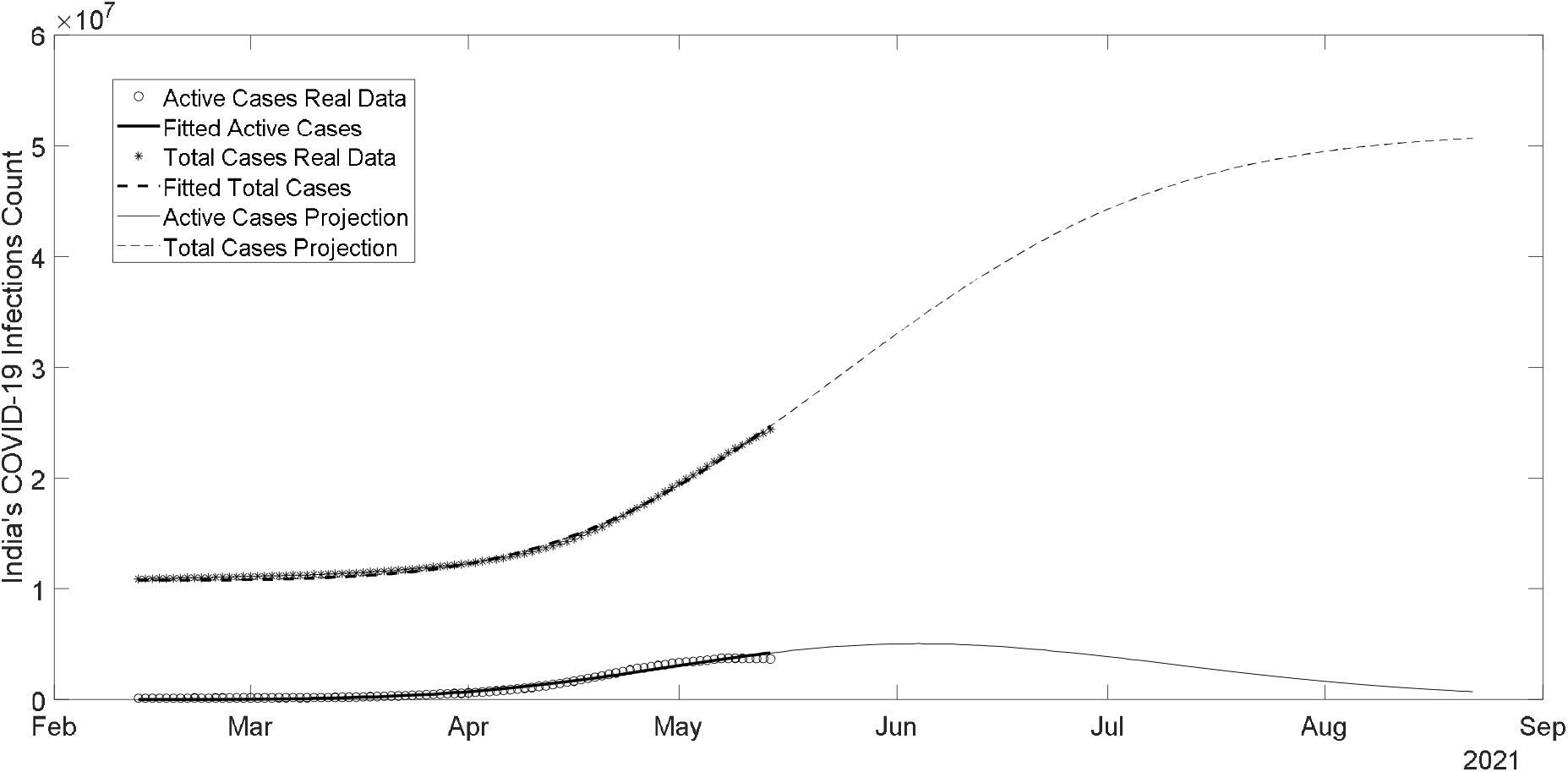
Comparison of the fitted values of active and total cases with the actual values and future prediction for India based on the data from 13^th^ February to 14^th^ May 2021. The parameters for this fit are: a = 0.236576657262; b = 0.008817517656; *I*(0) = 4806; *µ* = 0. 088195225005.

A fit of the data from 13^th^ February to 14^th^ May 2021 gives the parameters as a = 0.236576657262; b = 0.008817517656; *I*(0) = 4806; *µ* = 0.088195225005. Notice that parameter *b* is positive for the first time since 13^th^ February 2021, indicating a decrease in the infection rate.

According to [4], a point of inflection on the curve (1) of actively infected cases where its decline starts is given by

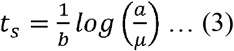

The parameters for the final fit give *t*_*s*_ as equal to 111.9, which indicates the decline of active cases may start from 4^th^ June 2021.

## Data Availability

All the data used in the study is available at
https://www.worldometers.info/coronavirus/country/india. (May 15, 2021)

https://www.worldometers.info

